# Clinical and Genetic Characteristics of Covid-19 Patients from UK Biobank

**DOI:** 10.1101/2020.05.05.20075507

**Authors:** David A. Kolin, Scott Kulm, Olivier Elemento

## Abstract

**Objective:** To explore both clinical and genetic risk factors for Covid-19 in a cohort from the United Kingdom.

**Design:** Prospective cohort study.

**Participants:** 669 positive Covid-19 patients within a cohort of 502,536 UK Biobank participants, recruited between 2006 and 2010.

**Main Outcome Measures:** The main outcome measure was Covid-19 positive status, determined by the presence of any positive test for a single individual. We also assessed risk factors for inpatient and outpatient status for Covid-19 positive individuals.

**Results:** We found that black participants were at over three times increased risk of testing positive for Covid-19, relative to white participants, even after adjusting for confounders (adjusted relative risk [ARR] 3.14, 95% confidence interval [CI] 2.28 to 4.31). Asian participants were also at higher risk of Covid-19 (ARR 2.03, 95% CI 1.40 to 2.95). Next, we analyzed the association of comorbidities with Covid-19. We found that participants were at increased risk of Covid-19 if they had chronic obstructive pulmonary disease (ARR 1.54, 95% CI 1.02 to 2.31) or ischemic heart disease (ARR 1.56, 95% CI 1.18 to 2.07). However, there was no evidence that either angiotensin converting enzyme inhibitors (ARR 1.32, 95% CI 0.95 to 1.84) or angiotensin II receptor blockers (ARR 1.37, 95% CI 0.94 to 1.98) increased the risk of Covid-19. We confirmed that blood type A was associated with Covid-19 relative to blood type O individuals, and we also found that the HLA variant DQA1_509 was enriched in Covid-19 positive cases, even after Bonferroni correction (P = 1.0 x 10^−5^).

**Conclusions:** In this study, we found that black and Asian participants were at increased risk of Covid-19, even after adjusting for confounders. We also identified a novel genetic association with the HLA variant DQA1_509. Further investigations of genetic associations with Covid-19 may lead to important discoveries of genetic drivers of severe disease.

## Introduction

The coronavirus disease 2019 (Covid-19) pandemic has afflicted hundreds of thousands of people worldwide. Evidence suggests that minorities are at increased risk of Covid-19. In the United States, the Center for Disease Prevention and Control found that 33% of people hospitalized with Covid-19 are African American, even though African Americans make up only 13% of the U.S. population.^1^ The risk of death from Covid-19 for black Americans is 92.3 per 100,000 individuals, while for white Americans the risk of death is 45.2 per 100,000 individuals.

Similarly, in the United Kingdom evidence suggests that Covid-19 is devastating minority communities. Approximately one-third of Covid-19 patients admitted to 201 critical care units in the United Kingdom were ethnic minorities.^2^ However, limitations in data collection make tracking mortality amongst different minorities difficult. Many countries – including the United Kingdom – do not require incorporating demographic data like ethnicity when reporting mortality.

There has also been considerable interest in better understanding the effects of medication use on Covid-19 susceptibility. Because coronaviruses use angiotensin converting enzyme 2 to gain entry into cells, concerns arose regarding the use of medications targeting the angiotensin system in Covid-19 patients. However, some recent evidence suggests that angiotensin converting enzyme inhibitors and angiotensin II receptor blockers do not increase the risk of Covid-19.^3,4^

In order to better understand risk factors, including demographics and medication use, we analyzed data from 669 Covid-19 positive patients from UK Biobank.

## Methods

This prospective cohort study includes a population of 502,536 participants between the ages of 40 and 69 recruited from the United Kingdom from 2006 to 2010. Recently, Public Health England began providing UK Biobank with information on participants’ Covid-19 test results from March 16, 2020 onwards. Samples obtained from patients were kept on a medium salt solution during transfer to a testing facility, where samples were tested for Covid-19 using polymerase chain reaction. Approximately four days elapsed between the time of sample retrieval and the time of data transfer into the Public Health England system. We used frequencies, percentages, and relative risk [RR] estimates to characterize associations. For continuous variables, relative risk was calculated using quasi-Poisson models. Adjusted relative risk [ARR] estimates accounted for age, sex, body-mass index, systolic blood pressure, race, and Townsend deprivation index.

## Results

Among 502,536 participants, 1,474 (0.3%) patients were tested for Covid-19, of which 1,116 (75.7%) were inpatients. The cohort underwent 2,724 tests, with a total of 1,160 (42.6%) positive tests. One participant was tested 20 times. Although, Public Health England notes that duplicate tests may be present in this study due to the arrival of results via several different routes. The majority of tests were upper respiratory tract swabs (33.0%), nasal swabs (17.6%), or throat swabs (13.2%). Of the patients tested, 669 (45.4%) tested positive at least one time (Table 1). The mean age of Covid-19 positive participants was 57.5 years, 56.5% were male, and mean body-mass index was 29.2.

**Table 1.** Baseline Characteristics of UK Biobank Participants

| **Characteristic** | **All**<br>**Participants**<br>**(N = 502536)** | **All Tests**<br>**Negative**<br>**(N = 805)** | **At Least**<br>**One Positive**<br>**Test**<br>**(N = 669)** |
| --- | --- | --- | --- |
| **Baseline and demographic** |  |  |  |
| Mean age, s.d. (years) | 56.5 (8.1) | 57.9 (8.7) | 57.5 (8.7) |
| Male, no. (%) | 229134 (45.6) | 409 (50.8) | 378 (56.5) |
| Mean body-mass index, s.d.* | 27.4 (4.8) | 28.3 (5.7) | 29.2 (5.5) |
| Mean systolic blood pressure, s.d. (mmHg) | 139.7 (19.7) | 140.1 (20.6) | 141.3 (20.4) |
| Race, no. (%) |  |  |  |
| White | 472725 (94.1) | 741 (92.0) | 565 (84.5) |
| Asian | 12287 (2.4) | 22 (2.7) | 33 (4.9) |
| Black | 9079 (1.8) | 23 (2.9) | 48 (7.2) |
| Mean Townsend deprivation index, s.d. | -1.3 (3.1) | -0.3 (3.5) | 0.0 (3.6) |
| **Social habit** |  |  |  |
| Smoking, no. (%) |  |  |  |
| Never | 273537 (54.4) | 354 (44.0) | 303 (45.3) |
| Previous | 173070 (34.4) | 307 (38.1) | 285 (42.6) |
| Current | 52979 (10.5) | 141 (17.5) | 72 (10.8) |
| Alcohol use, no. (%) |  |  |  |
| Never | 40648 (8.1) | 85 (10.6) | 84 (12.6) |
| Once or twice a week | 129297 (25.7) | 191 (23.7) | 157 (23.5) |
| Three or four times a week | 115445 (23.0) | 159 (19.8) | 128 (19.1) |
| Daily or almost daily | 101774 (20.3) | 168 (20.9) | 113 (16.9) |
| **Comorbidity** |  |  |  |
| Cancer, no. (%) | 41700 (8.3) | 84 (10.4) | 58 (8.7) |
| Diabetes, no. (%) | 26402 (5.3) | 71 (8.8) | 66 (9.9) |
| Chronic obstructive pulmonary disease, no. (%)<sup>1</sup> | 11823 (2.4) | 43 (5.3) | 28 (4.2) |
| Asthma, no. (%) | 58278 (11.6) | 111 (13.8) | 102 (15.2) |
| Ischemic heart disease, no. (%)<sup>2</sup> | 22727 (4.5) | 79 (9.8) | 58 (8.7) |
| Hypothyroidism, no. (%) | 24240 (4.8) | 33 (4.1) | 30 (4.5) |
| Hypercholesterolemia, no. (%) | 61635 (12.3) | 142 (17.6) | 111 (16.6) |
| Allergic rhinitis, no. (%) | 28148 (5.6) | 40 (5.0) | 42 (6.3) |
| Depression, no. (%) | 28208 (5.6) | 75 (9.3) | 56 (8.4) |
| **Serology** |  |  |  |
| Mean white blood cell count, s.d. | 6.9 (2.1) | 7.2 (2.2) | 7.4 (5.2) |
| Mean red blood cell count, s.d. | 4.5 (0.42) | 4.5 (0.45) | 4.6 (0.5) |
| Mean hemoglobin concentration, s.d. | 14.2 (1.2) | 14.1 (1.3) | 14.3 (1.5) |
| Mean corpuscular volume, s.d. | 91.1 (4.6) | 91.3 (5.1) | 90.8 (5.1) |
| Mean corpuscular hemoglobin concentration, s.d. | 34.5 (1.1) | 34.4 (1.0) | 34.4 (1.0) |
| Mean platelet count, s.d. | 253.0 (60.0) | 250.1 (61.1) | 249.5 (64.7) |
| Mean lymphocyte count, s.d. | 2.0 (1.2) | 2.0 (1.2) | 2.3 (4.8) |
| Mean monocyte count, s.d. | 0.5 (0.3) | 0.5 (0.2) | 0.5 (0.4) |
| Mean neutrophil count, s.d. | 4.2 (1.4) | 4.5 (1.5) | 4.4 (1.5) |

\*Body-mass index is the weight in kilograms divided by the square of the height in meters.
<sup>1</sup>Chronic obstructive pulmonary disease was defined as a diagnosis of emphysema and/or bronchitis.
<sup>2</sup>Ischemic heart disease was categorized as history of myocardial infarction or angina.

Black participants were disproportionately Covid-19 positive (supplementary eTable 1). Compared to white participants, black participants were at over four times increased risk of testing positive for Covid-19 (RR 4.35, 95% confidence interval [CI] 3.24 to 5.83). Amongst the 48 black participants who tested positive for Covid-19, 8 (16.7%) had diabetes. In exploratory analyses adjusting for age, sex, body-mass index, Townsend deprivation score, and history of diabetes, angina, or myocardial infarction, black participants remained at increased risk of Covid-19 (ARR 3.14, 95% CI 2.28 to 4.31). Asian participants were also at increased risk of testing positive for Covid-19 compared to white participants (ARR 2.03, 95% CI 1.40 to 2.95). Analyses of several serological markers identified that baseline white blood cell count, lymphocyte count, and monocyte count were associated with Covid-19.

Next, we investigated the association of common classes of medications with Covid-19 (Table 2). Participants who used angiotensin converting enzyme inhibitors were at increased risk of Covid-19 (RR 1.76, 95% CI 1.28 to 2.41) (supplementary eTable 2). However, adjustment reduced the strength of the association (ARR 1.32, 95% CI 0.95 to 1.84). Participants using angiotensin receptor blockers had a similar adjusted risk ratio (ARR 1.37, 95% CI 0.94 to 1.98). Of the 48 black Covid-19 patients, 11 (22.9%) were using either an angiotensin converting enzyme inhibitor or an angiotensin II receptor blocker. Of all black participants, 611 (6.7%) were using either of the two medications. Adjusted risk ratios for use of non-steroidal anti-inflammatory drugs and acetaminophen were 1.02 (95% CI 0.86 to 1.22) and 1.25 (95% CI 1.03 to 1.51), respectively. Analyses of Covid-19 positive inpatients and outpatients found that the groups were similar across baseline characteristics and medication use (supplementary eTable 3 to 6).

**Table 2.** Frequency of Medication Use and Detection of Covid-19

| **Medication Class** | **All Participants**<br>**(N = 502536)** | **All Tests Negative**<br>**(N = 805)** | **At Least One Positive Test**<br>**(N = 669)** |
| --- | --- | --- | --- |
| Non-steroidal anti-inflammatory drug, no. (%)<sup>1</sup> | 133030 (26.5) | 261 (32.4) | 196 (29.3) |
| Angiotensin converting enzyme inhibitor, no. (%)<sup>2</sup> | 18623 (3.7) | 45 (5.6) | 41 (6.1) |
| Angiotensin II receptor blocker, no. (%)<sup>3</sup> | 5497 (1.1) | 16 (2.0) | 14 (2.1) |
| Dihydropyridine calcium channel blocker, no. (%)<sup>4</sup> | 26832 (5.3) | 80 (9.9) | 65 (9.7) |
| Beta blocker, no. (%)<sup>5</sup> | 31725 (6.3) | 79 (9.8) | 65 (9.7) |
| Thiazolidinedione, no. (%)<sup>6</sup> | 1838 (0.4) | 5 (0.6) | 4 (0.6) |
| Sulfonylurea, no. (%)<sup>7</sup> | 4923 (1.0) | 21 (2.6) | 17 (2.5) |
| **Other Common Therapies** |  |  |  |
| Acetaminophen, no. (%) | 93668 (18.6) | 202 (25.1) | 154 (23.0) |
| Levothyroxine, no. (%) | 20344 (4.0) | 32 (4.0) | 27 (4.0) |
| Metformin, no. (%) | 14179 (2.8) | 42 (5.2) | 43 (6.4) |
| Glucosamine, no. (%) | 32135 (6.4) | 37 (4.6) | 32 (4.8) |
| Cod liver oil capsule, no. (%) | 27963 (5.6) | 37 (4.6) | 30 (4.5) |

<sup>1</sup>Non-steroidal anti-inflammatory drugs included aspirin, ibuprofen, diclofenac, naproxen, indomethacin, celecoxib, and meloxicam.
<sup>2</sup>Angiotensin converting enzyme inhibitors included captopril, enalapril, lisinopril, fosinopril, ramipril, and quinapril.
<sup>3</sup>Angiotensin II receptor blockers included losartan, candesartan, eprosartan, irbesartan, olmesartan, telmisartan, and valsartan.
<sup>4</sup>Dihydropyridine calcium channel blockers included amlodipine, felodipine, isradipine, nicardipine, and nifedipine.
<sup>5</sup>Beta blockers included acebutolol, atenolol, bisoprolol, carvedilol, labetalol, metoprolol, nadolol, nebivolol, pindolol, and propranolol.
<sup>6</sup>Thiazolidinediones included rosiglitazone, troglitazone, and pioglitazone.
<sup>7</sup>Sulfonylureas included glipizide, glibenclamide, glibornuride, gliclazide, gliquidone, acetohexamide, tolbutamide, chlorpropamide, and tolazamide.

Lastly, we investigated the genetics of participants with Covid-19. ABO blood types of participants were inferred through their genetic profiles.^5^ Participants with blood type A had increased odds of at least one positive Covid-19 test relative to blood type O participants (P = 0.003), consistent with the previously-noted association (supplementary eTable 7).^6^ However, no significant associations were identified when comparing Covid-19 positive inpatients to positive outpatients. We then analyzed whether any polymorphisms in ACE2, TMPRSS2, or the HLA region were enriched in either Covid-19 positive individuals or in Covid-19 positive inpatients. We found that a single HLA variant (DQA1_509, P = 1.0 x 10^−5^) was enriched in Covid-19 positive cases, even after Bonferroni correction (eFigs 1 and 2). Systematic genome wide association studies were then conducted with adjustments for age, sex, and the first ten genetic principal components. No significant variants (P < 5×10^−8^) were identified when comparing participants with any positive Covid-19 test relative to those with no positive test (669 cases and 481,583 controls) or when comparing Covid-19 positive inpatients to positive outpatients (574 cases and 95 controls) (eFigs 3 and 4). Finally, polygenic risk scores for Covid-19 were unable to significantly stratify either Covid-19 positive individuals or the full cohort (eFigs 5 and 6).

## Discussion

Multiple reports both in the United States and the United Kingdom have shown that Covid-19 is disproportionately affecting black populations.^7^ In this cohort of study participants from the United Kingdom, we found that risk of Covid-19 was over fourfold higher for black participants than for white participants. Prior to this study, data fully adjusting for comorbidities in black populations had not been reported.^8^ Our adjusted effect estimates, accounting for a range of comorbidities, did not nullify the risk of Covid-19 in black participants. One possible explanation for the sustained increased risk, which we were unable to control for, was the number and duration of social interactions sustained by individuals, an important consideration given the highly contagious nature of Covid-19. This study lends some support to the reported hypothesis that upregulation of angiotensin-converting enzyme 2 is a risk factor for Covid-19; however, adjustment for several risk factors reduced the strength of the association.^9,10^ Differential effects of medications targeting the angiotensin system in black individuals may partially explain the increased prevalence of Covid-19 in black populations.^11^ However, additional data is required to support this conjecture. Finally, the analysis presented herein is one of the first genetic explorations of Covid-19 positive individuals. We identified a novel association in the HLA region, and we confirmed the association of Covid-19 with blood type A. Our findings suggest that genetic drivers of Covid-19 may emerge as additional data is released.

## Data Availability

UK Biobank is a cohort of over 500,000 patients that is accessible to all researchers via an application process.

## Ethical approval

This study was approved by Weill Cornell Medical College. The UK Biobank is overseen by an independent advisory committee, and the data used within this analysis was accessed through the approved application #47137. Because the UK Biobank uses de-identified patient data and uses an independent ethics committee, separate IRB approval is not necessary for UK Biobank projects.

## Contributors

DAK, SK, and OE conceived and designed the study. SK obtained access to the data. DAK and SK analyzed the data and drafted the manuscript’s initial version. All authors provided important insights during the data analysis. All authors had full access to the data, contributed to the interpretation of the data, and revised the manuscript. DAK is the guarantor. The corresponding author attests that all listed authors meet authorship criteria and that no others meeting the criteria have been omitted.

## Patient and Public Involvement

All details about patient and public involvement in the study can be found both within the manuscript and in-detail at the UK Biobank website: https://www.ukbiobank.ac.uk/. The findings of our report will be widely disseminated on social media platforms and in the press, allowing participants to fully engage with the disseminated study results.

## Data Sharing

All data used for this study can be found at https://www.ukbiobank.ac.uk/.

## Copyright/license for publication

The Corresponding Author has the right to grant on behalf of all authors and does grant on behalf of all authors, *a worldwide licence* to the Publishers and its licencees in perpetuity, in all forms, formats and media (whether known now or created in the future), to i) publish, reproduce, distribute, display and store the Contribution, ii) translate the Contribution into other languages, create adaptations, reprints, include within collections and create summaries, extracts and/or, abstracts of the Contribution, iii) create any other derivative work(s) based on the Contribution, iv) to exploit all subsidiary rights in the Contribution, v) the inclusion of electronic links from the Contribution to third party material where—ever it may be located; and, vi) licence any third party to do any or all of the above.

## Patient Consent

Each UK Biobank participant fully consented to participate in the UK Biobank study.

## References

1 COVID-19 in Racial and Ethnic Minority Groups. 2020.

2 Pareek M, Bangash MN, Pareek N, et al. Ethnicity and COVID-19: an urgent public health research priority. The Lancet 2020;395:1421–2. doi:10.1016/S0140-6736(20)30922-3

3 Mancia G, Rea F, Ludergnani M, et al. Renin–Angiotensin–Aldosterone System Blockers and the Risk of Covid-19. New England Journal of Medicine Published Online First: 1 May 2020. doi:10.1056/NEJMoa2006923

4 Mehra MR, Desai SS, Kuy S, et al. Cardiovascular Disease, Drug Therapy, and Mortality in Covid-19. New England Journal of Medicine Published Online First: 1 May 2020. doi:10.1056/NEJMoa2007621

5 Groot HE, Villegas Sierra LE, Said MA, et al. Genetically Determined ABO Blood Group and its Associations With Health and Disease. Arteriosclerosis, Thrombosis, and Vascular Biology 2020;40:830–8. doi:10.1161/ATVBAHA.119.313658

6 Zhao J, Yang Y, Huang H, et al. Relationship between the ABO Blood Group and the COVID-19 Susceptibility. Epidemiology 2020. doi:10.1101/2020.03.11.20031096

7 Khunti K. Is ethnicity linked to incidence or outcomes of covid-19? BMJ Published Online First: 20 April 2020. doi:https://doi.org/10.1136/bmj.m1548

8 Yancy CW. COVID-19 and African Americans. JAMA Published Online First: 15 April 2020. doi:10.1001/jama.2020.6548

9 Fang L, Karakiulakis G, Roth M. Are patients with hypertension and diabetes mellitus at increased risk for COVID-19 infection? The Lancet Respiratory Medicine 2020;8:e21. doi:10.1016/S2213-2600(20)30116-8

10 Tignanelli CJ, Ingraham NE, Sparks MA, et al. Antihypertensive drugs and risk of COVID-19? The Lancet Respiratory Medicine Published Online First: March 2020. doi:10.1016/S2213-2600(20)30153-3

11 Exner DV, Dries DL, Domanski MJ, et al. Lesser Response to Angiotensin-Converting–Enzyme Inhibitor Therapy in Black as Compared with White Patients with Left Ventricular Dysfunction. New England Journal of Medicine 2001;344:1351–7. doi:10.1056/NEJM200105033441802

